# Human genomics of the humoral immune response against polyomaviruses

**DOI:** 10.1101/2020.11.02.20224402

**Authors:** F. Hodel, A.Y. Chong, P. Scepanovic, Z.M. Xu, O. Naret, C.W. Thorball, S. Rüeger, P. Marques-Vidal, P. Vollenweider, M. Begemann, H. Ehrenreich, N. Brenner, N. Bender, T. Waterboer, A. J. Mentzer, A.V.S. Hill, C. Hammer, J. Fellay

## Abstract

Human polyomaviruses are widespread in human populations and are able to cause severe disease in immunocompromised individuals. There remains an incomplete understanding of the potential impact of human genetic variation on inter-individual responses to polyomaviruses.

To identify human genetic determinants of the humoral immune response against polyomaviruses, we performed genome-wide association studies and meta-analyses of qualitative and quantitative immunoglobulin G (IgG) responses against the major capsid protein VP1 of Human polyomavirus 6 (HPyV6), BK virus (BKPyV), JC virus (JCPyV), Merkel Cell Polyomavirus (MCPyV) and WU polyomavirus (WUPyV), in a total of 15,660 individuals of European ancestry from CoLaus, UK Biobank and GRAS, three independent studies.

We observed significant associations for all tested viruses: JCPyV, HPyV6 and MCPyV associated with HLA class II variation; BKPyV and JCPyV with variants in the *FUT2* gene, responsible for secretor status; MCPyV with variants in the *STING1* gene, involved in interferon induction; and WUPyV with a functional variant in the *MUC1* gene, previously associated with risk for gastric cancer.

These results provide insights into the genetic control of a family of very prevalent human viruses, highlighting genes and pathways that play a modulating role in human humoral immunity.

## Introduction

The variability of the humoral response to antigenic stimulation has been documented for a long time ^1^. While it is partly due to demographic and environmental factors, a substantial part of this variation is heritable and can thus be attributed to differences in the host genome. For example, human genetic factors were estimated to account for 35% (±6%) and 21% (±5%) of the variance in *H. pylori* and *T. gondii* antibody levels, and 60% (±11%) and 57% (±11%) of the variance in cytomegalovirus (CMV) and Epstein–Barr virus (EBV) serostatus, respectively ^2^. This high heritability makes serological phenotypes highly promising targets for human genomic studies. Indeed, specific genetic variants have already been associated with variation in immunoglobulin G (IgG) levels. Most variants identified to date are located within the major histocompatibility complex (MHC) locus on chromosome 6 ^3–5^. This highly polymorphic complex notably encodes the class I and class II human leucocyte antigen (HLA) proteins, which present antigenic epitopes to effector cells of the immune system - a key role at the crossroad between innate and acquired immune responses.

Polyomaviruses are small (40–50 nm), double-stranded DNA viruses belonging to the *Polyomaviridae* family, with an average genome size of 5,000 base pairs ^6,7^. First discovered in rodents in the 1950s, the first human polyomavirus, namely JCPyV, was discovered in 1965 in a patient with progressive multifocal leukoencephalopathy (PML) ^8,9^. Serological surveys indicate that human polyomaviruses are ubiquitous, with most of the world’s population infected by several of them during childhood ^10^. In most cases, polyomavirus infections are asymptomatic. After acute infection, they silently persist in healthy individuals. However, they can lead to symptoms and even severe disease in immunocompromised individuals. As an example, reactivation of JC polyomavirus (JCPyV) can cause PML in patients with low CD4^+^ T cell counts, such as individuals with AIDS or on chronic immunosuppressive therapy, and BK polyomavirus (BKPyV) is known to cause renal disease in kidney transplanted patients ^6,7,11,12^.

A better understanding of the genetic control of the human immune response against polyomaviruses may provide new insights into pathologic mechanisms. We therefore searched for associations between human genetic variation and both differences in IgG levels and serostatus against five human polyomaviruses: BKPyV, JCPyV, Human polyomavirus 6 (HPyV6), Merkel Cell Polyomavirus (MCPyV), and WU polyomavirus (WUPyV), using genome-wide association study (GWAS) and meta-analysis in a total of 15,660 healthy individuals of European ancestry from three studies from Switzerland, the United Kingdom and Germany.

## Materials and methods

### 1. Study cohorts

#### a. The CoLaus study

The *Cohorte Lausannoise* (CoLaus) is a population-based prospective study that started in 2003 in Lausanne, Switzerland ^13^. It includes 6,188 individuals of European descent (47.5% male) initially aged 35 to 75 years (mean ± SD: 51.1 ± 10.9), who were randomly selected from the general population and continuously undergo a follow-up every 5 years. Detailed phenotypic information was obtained from every study participant through questionnaires, a physical assessment and biological measurement of blood and urine markers. The institutional Ethics Committee of the University of Lausanne, which afterwards became the Ethics Commission of Canton Vaud (www.cer-vd.ch) approved the baseline CoLaus study (reference 16/03, decisions of 13th January and 10th February 2003), and written consent was obtained from all participants.

#### b. The GRAS Data Collection

The Göttingen Research Association for Schizophrenia (GRAS) Data Collection consists of 2,363 immunocompetent adults of European ancestry, comprising 1,147 anonymized blood donors (62.0% male, mean age ± SD: 37.5 ± 13.2) and 1,216 individuals with psychiatric diagnoses (64.9% male, mean age ± SD: 40.6 ± 13.5) ^14,15^. All study participants provided informed consent, including genetic testing, and the GRAS Data Collection has been approved by the ethical committee of the Georg-August-University of Göttingen (master committee) as well as by the respective local regulatory / ethical committees of all collaborating centers ^15^.

#### c. The UK Biobank

The UK Biobank (UKB) is a population-based prospective study whose recruitment process has been described previously ^16^. Briefly, half a million men and women aged 40-69 years (45.6% male, mean age ± SD: 56.5 ± 8.1) attended one of 22 UKB assessment centers located throughout England, Scotland and Wales between 2006 and 2010. All participants completed a touchscreen questionnaire, verbal interview and had a range of physical measurements and blood, urine and saliva samples taken for long-term storage. A subset of 20,000 individuals attended a repeat assessment between 2012 and 2013. This project was conducted with approved access to UKB data under application numbers 50085 (PI: Fellay) and 43920. All UKB participants provided informed consent at recruitment. Ethics approval for the UKB study was obtained from the North West Centre for Research Ethics Committee (11/NW/0382).

### 2. Serological analyses

To assess humoral responses to polyomaviruses, serum samples were independently shipped by the three studies to the Infections and Cancer Epidemiology Division at the German Cancer Research Center (Deutsches Krebsforschungszentrum, DKFZ) in Heidelberg. Seroreactivity against BKPyV, HPyV6, JCPyV, MCPyV and WUPyV viral capsid protein 1 (VP1) was measured at serum dilution 1:1000 using multiplex serology based on glutathione-S-transferase (GST)-VP1 fusion capture immunosorbent assays combined with fluorescent bead technology ^17^. Antigen preparation and methodology, as well as robustness of seroprevalence estimates, have been described previously for human polyomaviruses ^18^. For the quantitative IgG level association analyses including only seropositive individuals, results above assay cut-off of 250 mean fluorescence intensity (MFI) were used. For the case-control study design, results were expressed as a binary output (serostatus: anti-polyomavirus IgG positive or negative) based on the same assay cut-off. Using the same assay, seroreactivity against six *H. pylori* antigens (HP1564 OMP, HP10 GroEL, HP547 CagA, HP887 VacA, HP73 UreaseA and HP875 Catalase) and four EBV antigens (VCA p18, EBNA, Zebra and EA-D) were measured in CoLaus.

### 3. DNA genotyping and statistical analyses

#### a. The CoLaus study

DNA samples from 5,399 CoLaus participants were genotyped for 799,653 single nucleotide polymorphisms (SNPs) using the BB2 GSK-customized Affymetrix Axiom Biobank array. Quality control procedures described by Anderson *et al*. were applied ^19^. A total of 4,781 individuals were included for further analyses. Variants were excluded if they were missing in >5% of the subjects or if they deviated significantly from Hardy-Weinberg equilibrium (HWE, P<10e-7), resulting in 697,923 SNPs kept for phasing and imputation.

To statistically estimate the haplotypes, the genotypes were phased using EAGLE2 v2.0.5 software and the Haplotype Reference Consortium (HRC) r1.1 reference panel ^20,21^. Genotype imputation was performed using two independent reference panels: the HRC reference panel and the merged 1000 Genomes Phase 3 and UK10K reference panel ^20,22,23^. Both phasing and imputation were performed using the Sanger Imputation Service (https://imputation.sanger.ac.uk). The imputed data set (N=89 million SNPs) consisted of a) SNPs imputed with HRC reference panel only, b) SNPs imputed with the merged reference panel only, and c) SNPs with the highest imputation information (INFO) score if imputed with both reference panels. To filter out poorly imputed or rare genotypes, SNPs with INFO score < 0.8, minor allele frequency (MAF) < 1% or significant deviation from HWE (P<10e-7) were excluded, resulting in a final number of 9,031,264 SNPs available for association analysis. We also imputed 111 four-digit classical HLA alleles using SNP2HLA and the T1DGC Immunochip/HLA reference panel ^24^.

Genome-wide association analyses were performed using mixed models in GCTA ^25^. HLA association analyses were performed using generalized linear models in PLINK v2 ^26^. We assumed an additive model for SNP associations and a dominant model for associations with HLA alleles. The top three principal components of ancestry, sex and age were used as covariates in all analyses.

#### b. The GRAS Data Collection

Genome-wide SNP genotyping of GRAS study participants was performed using an Axiom®myDesignTM genotyping array (Affymetrix, Santa Clara, CA, USA). The quality control steps have been described previously ^3^. Imputation of unobserved genotypes was performed using the 1,000 Genomes Project Phase 1 v3 haplotypes as reference panel. Genotypes were pre-phased with MaCH v19 and subsequently imputed by minimac ^27^. SNPs with a reported INFO score < 0.8 or a MAF < 5%, as well as markers on sex chromosomes, were excluded from downstream analyses. SNPs were also filtered on the basis of missingness (excluded if <95% genotyping rate), and marked deviation from HWE (excluded if P<5e-07). We then used linear regression models in PLINK v1.9 to test for association between ∼6 million SNPs and IgG responses using a continuous, quantitative approach (log-normalized IgG levels in seropositive samples) ^26^. The first three principal components of ancestry, calculated using GCTA (v1.24), as well as sex and age, were included as covariates in all analyses ^25^.

#### c. The UK Biobank

Genotyping and imputation of UK Biobank individuals have been fully described by Bycroft *et al*. ^28^. Briefly, samples were genotyped on either the UK BiLEVE Axiom array (Affymetrix) or UK Biobank Axiom array (Applied Biosystems). Genotypes were phased using SHAPEIT3 and the 1000 Genome phase 3 dataset as a reference, then imputed using IMUTE4 using the Haplotype Reference Consortium data, 1000 Genomes phase 3, and UK10K data as references. Serological markers of infection were measured for a random subset of 9,611 unrelated UKB participants (44.0% male, mean age ± SD: 56.5 ± 8.2) ^29^. Association analyses were carried out using rank-based inverse normal transformed MFI values and linear mixed models as implemented in BOLT-LMM ^30,31^, using age at recruitment and genetic sex as covariates, and variants with a MAF > 0.01 and an INFO score > 0.3.

### 4. Meta-analyses

Meta-analyses of the GWAS results obtained in the different cohorts were performed using a fixed-effect model in METAL ^32^. For BKPyV and JCPyV, summary statistics of all three studies were used. For HPyV6 and WUPyV only the results of CoLaus and GRAS studies were used, and for MCPyV only the results of UKB and GRAS were used, as the data were not available in the UKB and CoLaus, respectively (**Figure 1**). The meta-analyses followed the sample size-based strategy, considering P-value, sample size and direction of effect. Bonferroni-corrected significance level threshold of P=5.6e-09 (P=5e-08 divided by the eight GWAS performed) was applied. Similarly, for HLA alleles, we determined significance thresholds correcting for the number of alleles, the viruses tested, and the two study designs (P=4.3e-05).

**Figure 1.**
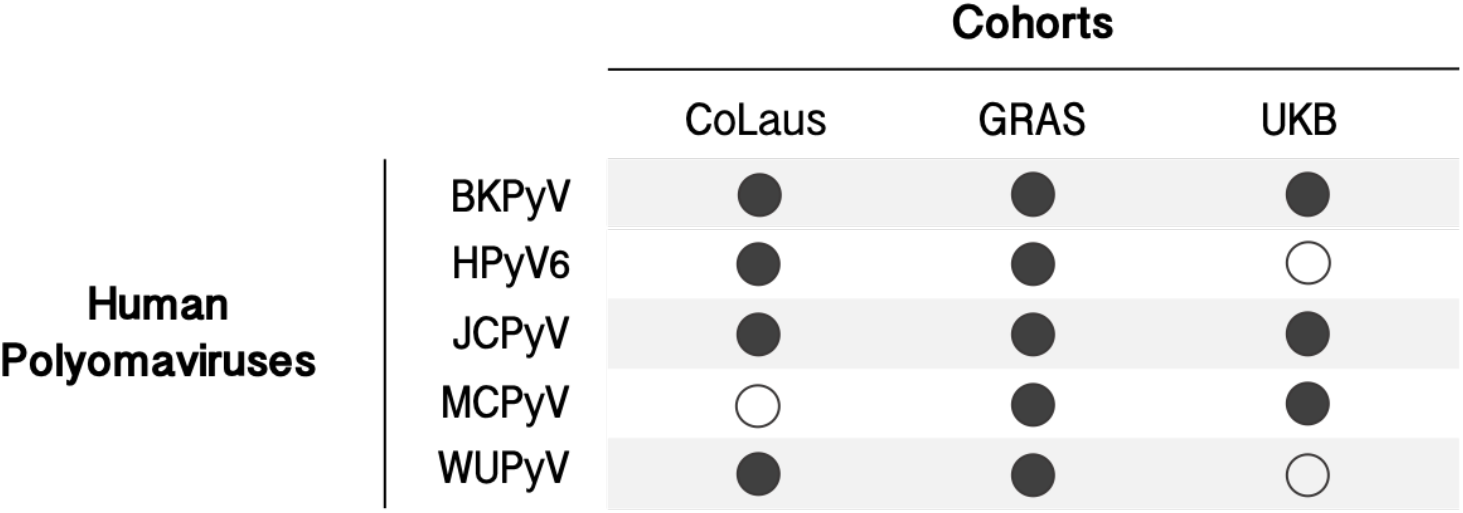
Visual representation of study data. Diagram representing data availability for each human polyomavirus by cohort. Black dots indicate that serological data are available in the cohort, white dots indicate that data are not available.

### 5. Inference of FUT2 secretor status

FUT2 secretor status was determined from the rs601338 genotype, where the wild-type rs601338 (G) encodes the secretor allele, whereas A is the nonsense “non-secretor” allele. Individuals with homozygous major (GG) or heterozygous (GA) genotypes were considered as secretors, and compared to homozygous minor (AA) non-secretors ^33^.

### 6. MUC1 full-length mRNA and Nanopore sequencing

A total of 12 RNA samples from gastrointestinal tissue (200 ng to 225 ng each) were obtained from the Genotype-tissue expression project (GTEx) ^34^. Samples were selected based on expression values of the MUC1 gene in the GTEx expression dataset and based on the genotype of the associated variant. Nine donors were male and three donors were female. Retrotranscription was performed using SMARTer™ PCR cDNA Synthesis Kit from Clonetech. We used the following forward and reverse primers to amplify the cDNA of the MUC1 transcript: 5’-GCGCCTGCCTGAATCTGTTC-3’ and 5’-CCCACATGAGCTTCCACACAC-3’. For the PCR reaction, we used LongAmp™ Taq 2X Master Mix (SuperScript from Life Technologies / Q5 High-Fidelity from New England BioLabs) and the following protocol: 30 seconds at 94°C for initial denaturation, following by 30 cycles at 94°C for 10 seconds, 55°C for 60 seconds, 65°C for 50 seconds and final extension at 65°C for 10 minutes. To pool the 12 samples for sequencing, we used native EXP-PBC001 barcoding kit on R9.4 flow cell. The following adapter sequences for MinION sequencing were added to the forward and reverse MUC1 primers, respectively: 5’-TTTCTGTTGGTGCTGATATT-3’ and 5’-ACTTGCCTGTCGCTCTATCTTC-3’. We used the SQK-MAP006 protocol for library preparation for nanopore sequencing on MinION (Oxford Nanopore Technologies, ONT). Briefly, the combined end repairing and dA-tailing step was performed using the NEBNext Ultra™ II End Repair/dA-Tailing Module (New England BioLabs). Samples were incubated for 5 min at 20°C followed by 5 min at 65°C. The reactions were purified by using Agencourt AMPure XP beads (Beckman Coulter) and further ligated to HP-adapter using Blunt/TA Ligase Master Mix (New England BioLabs) with 10 min incubation at room temperature. Adapted DNA was purified using Dynabeads MyOne Streptavidin C1 (Life Technologies) and washed to remove unbounded DNA. The captured cDNAlibrary was eluted by resuspending the beads in ONT’s Elution Buffer for 10 min at 37°C and the beads were pelleted using a magnetic rack leaving the supernatant containing the library. We used poretools ^35^ to extract fasta sequences from the ONT fast5 format, with a minimum length of 600 base pairs and forward and reverse strand sequence (2d reads). We then used nanocorrect ^36^ for computational error correction and aligned all corrected reads to a list of MUC1 isoforms using the LAST aligner ^37^. Each read was assigned to the isoform with the best alignment score.

## Results

### Seroprevalence of polyomaviruses

We included 3,689 individuals from CoLaus study (46% males, mean age ± SD: 53.1 ± 10.6), 2,363 from GRAS Data Collection (63% males, mean age ± SD: 39.1 ± 13.5) and 9,351 from the UKB (44% males, mean age ± SD: 56.5 ± 8.2), resulting in a total of 15,660 individuals with available genotyping data, serological data and covariates. All tested polyomaviruses had high seroprevalence in the three samples, with highest observed seroprevalence for WUPyV (96%), followed by BKPyV (91%), HPyV6 (84%), MCPyV (73%) and JCPyV (55%) (**Table 1**). In all cohorts combined, the serological responses against several polyomaviruses correlated with age and sex, confirming previous observations ^3,38,39^. We observed a significant decrease with age in antibody levels against BKPyV (P<2e-16), JCPyV (P=4.56e-03), MCPyV (P=5.77e-03) and WUPyV (P<2e-16); and lower IgG levels against HPyV6 (P=3.51e-15), JCPyV (P<2e-16) and MCPyV (P=1.07e-05) in female participants.

**Table 1.** Descriptive statistics of polyomavirus infections in the three cohorts.

|  | **CoLaus**<br>**(N = 3,689)** |  | **GRAS**<br>**(N = 2,363)** |  | **UKB**<br>**(N = 9,351)** |  |
| --- | --- | --- | --- | --- | --- | --- |
| | N seropositives | IgG level in seropositives (log, mean ± SD) | N seropositives | IgG level in seropositives (log, mean ± SD) | N seropositives | IgG level in seropositives (log, mean ± SD) |
| **BKPyV** | 3,112 (84%) | 7.63 ± 0.99 | 2,226 (94%) | 8.45 ± 0.98 | 8,911 (95%) | 7.96 ± 0.90 |
| **HPyV6** | 3,156 (86%) | 8.13 ± 0.88 | 1,938 (82%) | 8.30 ± 0.90 | - | - |
| **JCPyV** | 1,915 (52%) | 7.16 ± 0.90 | 1,268 (54%) | 7.15 ± 0.82 | 5,384 (58%) | 6.75 ± 0.88 |
| **MCPyV** | - | - | 1,871 (79%) | 8.01 ± 1.02 | 6,220 (67%) | 7.70 ± 1.01 |
| **WUPyV** | 3,536 (96%) | 7.85 ± 0.81 | 2,257 (96%) | 8.31 ± 0.81 | - | - |

- : not measured

### Genome-wide association studies and meta-analyses

We performed a total of nine independent GWAS using either a case-control study design (serostatus: antibody-positive versus antibody-negative) or a continuous, quantitative approach (IgG levels in seropositive individuals) to search for human genetic determinants of the antibody response to BKPyV and JCPyV in the three cohorts, to HPyV6 and WUPyV in CoLaus and GRAS, and to MCPyV in GRAS and UKB (**Figure 1**). We did not run a case-control analysis for WUPyV because of the very high seroprevalence of that virus (96%). Meta-analyses were conducted using GWAS summary statistics. For each of the five phenotypes, meta-analytic inflation factors λ ranged from 0.99 to 1.01 across the tested cohorts, indicating proper control of population stratification (**Figure S1**). In total, we identified four genome-wide significant loci, mapping to the *HLA* (for HPyV6, JCPyV and MCPyV), *FUT2* (for BKPyV and JCPyV), *STING1* (for MCPyV) and *MUC1* (for WUPyV) regions (**Figure 2**).

**Figure 2.**
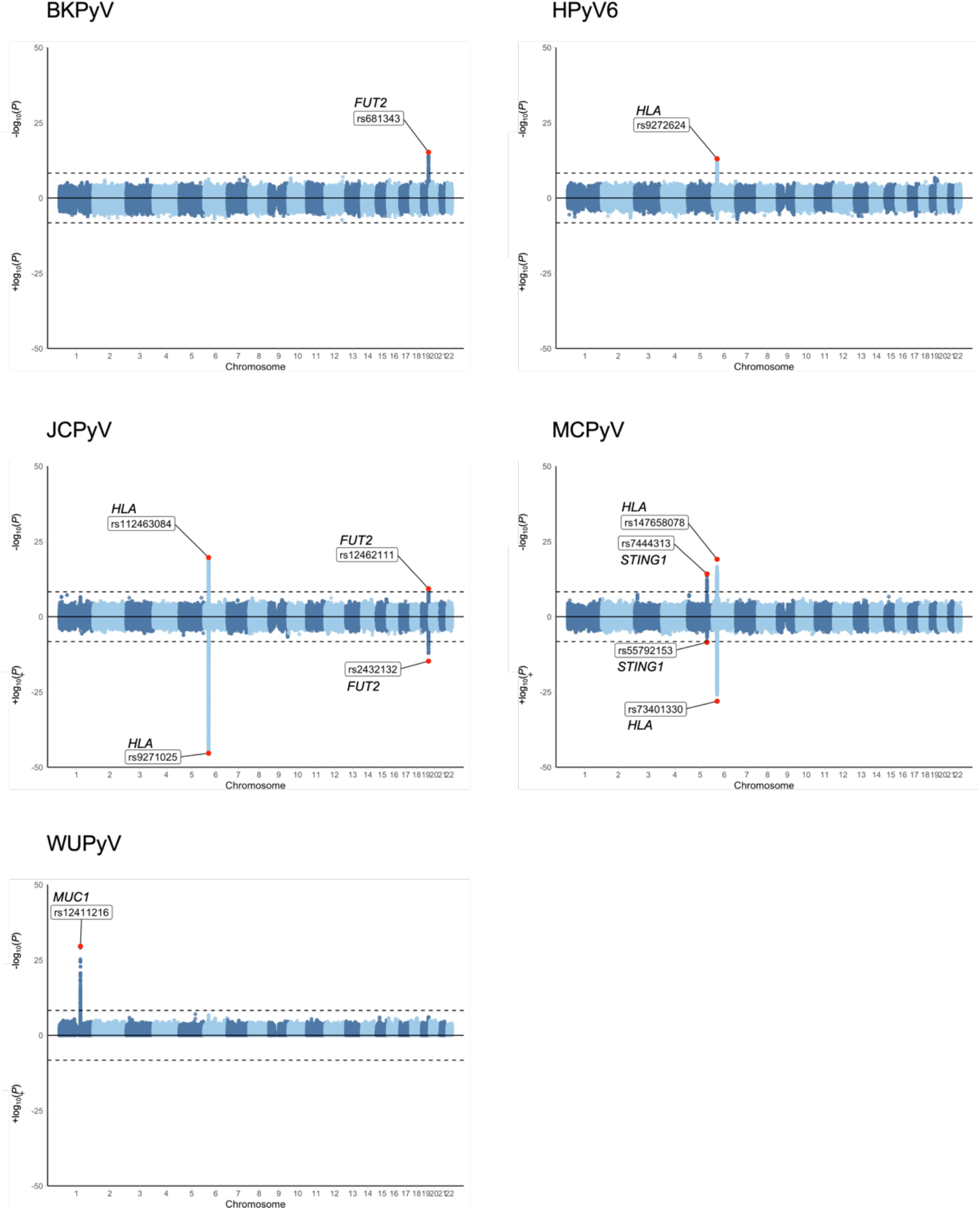
Chicago plots of genome-wide association results of the meta-analyses of antibody levels (top) and serostatus (bottom) against BKPyV, HPyV6, JCPyV, MCPyV and WUPyV. Chicago plots showing the significance of association of all SNPs across chromosomes 1 to 22. SNPs are plotted on the x-axis according to their physical position on each chromosome and the strength of the association with antibody levels is indicated on the y-axis (as −log10 P-value for IgG response, and as +log10 P-value for serostatus). The dashed line marks the Bonferroni-corrected genome-wide significance threshold of 5.6e-09. For each locus, the SNP with the most significant association is plotted in red and labeled. Serostatus was not tested for WUPyV due to very high seroprevalence. Chromosome X is not shown since data were not available for all studies.

### Role of HLA variation in antibody response to polyomaviruses

Significant associations were observed in the HLA class II region on chromosome 6 for HPyV6, JCPyV and MCPyV (**Figure 2**), with the lead variants being rs9272624 (P=9.62e-14), rs112463084 (P=2.18e-20) and rs147658078 (P=8.09e-20), respectively. To further dissect the association signals, we imputed 4-digit HLA alleles and tested them for association with anti-HPyV6, anti-JCPyV and anti-MCPyV VP1 IgG levels. The regional association plots are shown in **Figure S2** and resulting association p-values are listed in **Table 2**. We observed strong associations with quantitative seroreactivity for HLA-DRB1 and HLA-DQA1 alleles for all three human polyomaviruses. The size and directionality of the effects were virus-specific.

**Table 2.** Significant HLA association results.

| Antibody levels |  |  |  | Serostatus |  |  |  |
| --- | --- | --- | --- | --- | --- | --- | --- |
| Polyomavirus<br>(sample size) | Marker | Z-score | P-value | Polyomavirus<br>(sample size) | Marker | Z-score | P-value |
| HPyV6<br>(N=5,091) | DQA1*05:01 | -6.11 | 1.01e-09 | JCPyV<br>(N=15,400) | DQB1*06:02 | 15.97 | 2.08e-57 |
|  | DRB1*03:01 | -4.92 | 8.81e-07 |  | DQA1*01:02 | 15.39 | 2.07e-53 |
|  | DQB1*02:01 | -4.84 | 1.28e-06 |  | DRB1*15:01 | 14.13 | 2.58e-45 |
|  | DQA1*01:01 | 4.48 | 7.53e-06 |  | B*07:02 | 8.10 | 5.45e-16 |
|  | DRB1*01:01 | 4.33 | 1.47e-05 |  | C*07:02 | 6.92 | 4.69e-12 |
| JCPyV<br>(N=8,567) | DRB1*15:01 | 9.10 | 9.19e-20 |  | DRB1*13:01 | -6.27 | 3.71e-10 |
|  | DQB1*06:02 | 9.05 | 1.43e-19 |  | DQB1*06:03 | -5.97 | 2.43e-09 |
|  | DQA1*01:02 | 8.70 | 3.30e-18 |  | DQB1*02:01 | -5.87 | 4.27e-09 |
|  | DQB1*03:01 | -5.29 | 1.19e-07 |  | DRB1*03:01 | -5.65 | 1.57e-08 |
|  | B*07:02 | 4.46 | 8.12e-06 |  | DQA1*05:01 | -5.55 | 2.85e-08 |
|  | DRB1*03:01 | -4.21 | 2.57e-05 |  | DQB1*03:01 | -5.39 | 7.01e-08 |
|  | DQB1*02:01 | -4.16 | 3.17e-05 |  | DQA1*01:03 | -4.97 | 6.56e-07 |
|  | DQA1*05:01 | -4.14 | 3.51e-05 |  | B*08:01 | -4.97 | 6.70e-07 |
| MCPyV<br>(N=8,091) | DQA1*01:01 | -9.17 | 4.96e-20 |  | DQB1*05:02 | 4.88 | 1.07e-06 |
|  | DQB1*05:01 | -9.13 | 7.12e-20 |  | DRB1*16:01 | 4.74 | 2.13e-06 |
|  | A*29:02 | 8.64 | 5.85e-18 |  | DRB1*04:01 | -4.49 | 7.18e-06 |
|  | DRB1*15:01 | 8.58 | 9.68e-18 |  | C*07:01 | -4.44 | 8.94e-06 |
|  | DRB1*01:01 | -7.64 | 2.20e-14 | MCPyV<br>(N=11,714) | DQB1*06:02 | 13.65 | 1.91e-42 |
|  | DRB1*04:04 | 7.57 | 3.85e-14 |  | DRB1*15:01 | 13.54 | 9.35e-42 |
|  | DQB1*06:02 | 7.54 | 4.72e-14 |  | DQA1*01:02 | 11.28 | 1.59e-29 |
|  | DQA1*01:02 | 6.28 | 3.43e-10 |  | B*07:02 | 9.98 | 1.93e-23 |
|  | DRB1*11:01 | -5.92 | 3.24e-09 |  | C*07:02 | 8.49 | 2.12e-17 |
|  | C*07:02 | 5.63 | 1.86e-08 |  | DQA1*01:01 | -7.40 | 1.40e-13 |
|  | B*07:02 | 5.22 | 1.76e-07 |  | DQB1*05:01 | -7.12 | 1.07e-12 |
|  | C*16:01 | 4.79 | 1.64e-06 |  | DRB1*01:01 | -6.63 | 3.29e-11 |
|  | DQB1*03:01 | -4.61 | 4.12e-06 |  | DQA1*05:01 | -5.64 | 1.74e-08 |
|  | B*44:02 | -4.24 | 2.22e-05 |  | DRB1*11:01 | -5.57 | 2.60e-08 |
|  | DPB1*06:01 | 4.23 | 2.35e-05 |  | DRB1*04:04 | 5.40 | 6.86e-08 |
|  | B*44:03 | 4.18 | 2.95e-05 |  | DQB1*03:02 | 4.50 | 6.81e-06 |

### FUT2 non-secretor status is associated with stronger antibody responses to BKPyV and JCPyV

We observed significant associations in the *FUT2* gene on chromosome 19 for BKPyV and JCPyV (**Figure 2**). The fucosyltransferase 2 protein, encoded by *FUT2*, is expressed in epithelial cells and adds fucose to type 1 glycoprotein chains to form soluble ABH antigens that can be found in secretions and fluids ^33,40^. The top associated SNPs for BKPyV (rs681343, P=6.40e-16) and JCPyV (rs12462111, P=4.84e-10) are in high linkage disequilibrium (LD, r^2^ > 0.7) with the common *FUT2* nonsense variant rs601338 (P=6.81e-16 and P=1.89e-09, respectively). The homozygous genotype for the rs601338 minor allele (AA) is classically referred to as the “non-secretor” variant, as individuals homozygous for this null allele do not secrete blood group antigens at epithelial surfaces ^33,40^. Among the three cohorts, 24% of individuals were FUT2 non-secretors (AA), and 76% were FUT2 secretors (GG or GA). Overall, we observed that non-secretors had significantly higher IgG levels against both polyomaviruses in comparison to secretors (**Figure 3**).

**Figure 3.**
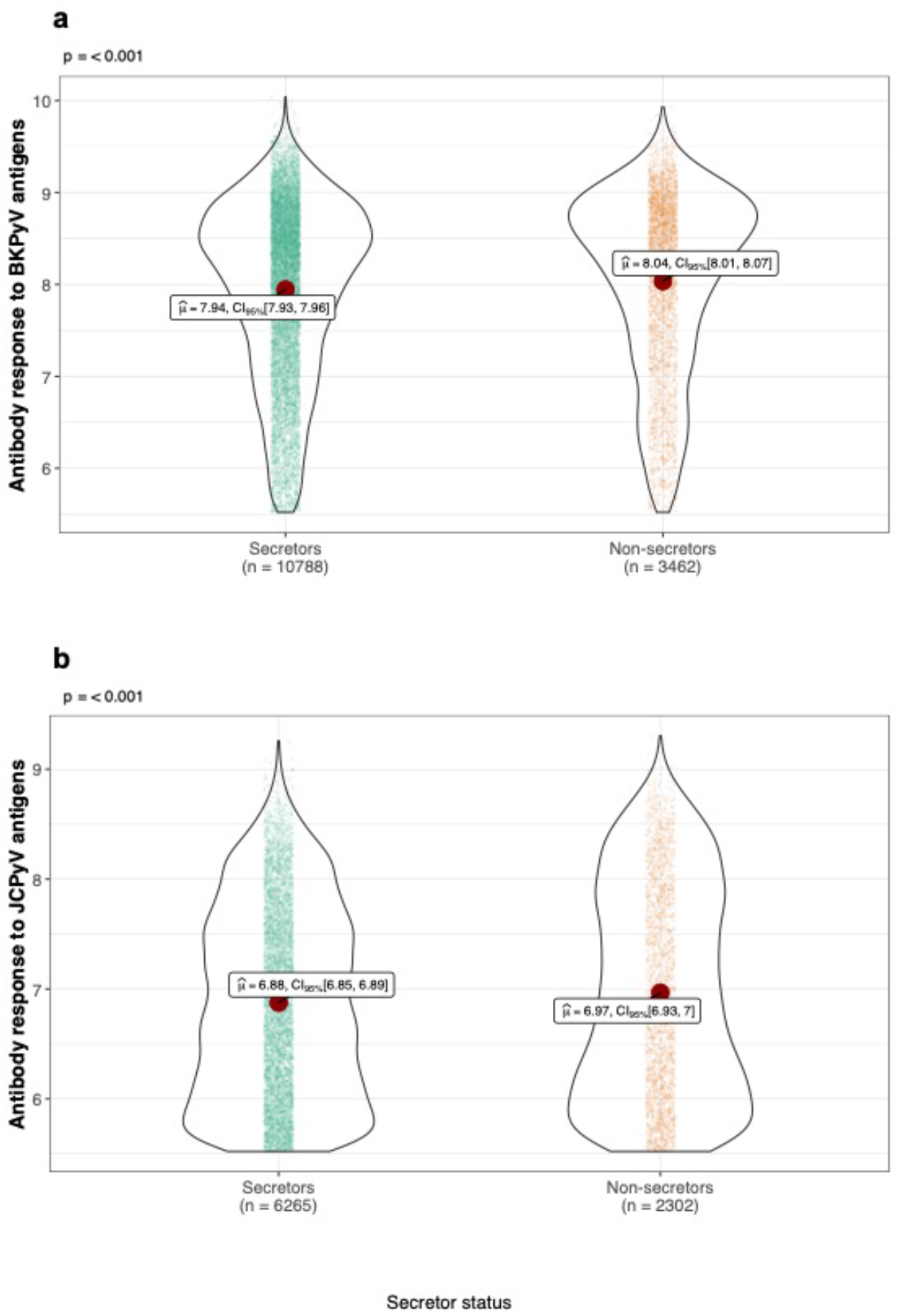
Serum immunoglobulin G (IgG) levels against BKPyV and JCPyV, by *FUT2* genotype / secretor status. Violin plots show the log-transformed antibody levels (in MFI) against a) BKPyV and b) JCPyV. Antibody levels are categorized according to individuals, FUT2 secretor or non-secretor genotype. Red dots represent the mean. P-value based on t-test are shown.

### Genomic variation in *STING1* is associated with antibody response to MCPyV

We observed genome-wide significant associations with anti-MCPyV IgG levels in the *STING1* gene on chromosome 5 (**Figure 2**). *STING1* encodes the endoplasmic reticulum resident membrane protein STING that functions as a major regulator of innate immunity. Upon viral infection and activation of systolic cyclic GMP-AMP synthase (cGAS), STING mediates type I interferon production by infected cells to protect them and neighboring cells from local infection ^41^. The top associated variant in meta-analysis using GRAS and UKB was rs7444313 (P=5.95e-15), a strong eQTL for *STING1* in the GTEx dataset, whose G allele was associated with a stronger IgG response against MCPyV VP1 ^34^.

### Alternative splicing of *MUC1* is associated with humoral immune response to WUPyV

We observed a genome-wide significant association with anti-WUPyV IgG levels on chromosome 1, mapping to the *MUC1* gene that encodes the Mucin1 protein (lead SNP rs12411216, P=2.70e-30; **Figure 2**). This variant is in perfect LD (r^2^=1) with the splicing variant rs4072037 (P=4.08e-30) located in the second exon of the *MUC1* gene. The rs4072037 T allele was associated with a stronger IgG response against WUPyV VP1. The same allele was previously associated with an increased risk of gastric cancer ^42,43^. Because infections with *H. pylori* and EBV have been implicated in the pathogenesis of gastric cancers, we searched for potential associations of rs4072037 with the humoral response to these pathogens ^44–47^. Using GWAS data available in CoLaus, we performed serological analyses for six different *H. pylori* antigens and four EBV antigens, but did not find evidence for a rs4072037-dependent differential antibody response (**Table S1**).

### Relative abundance of short MUC1 isoforms depends on rs4072037 allele status

The SNP rs4072037 has been functionally investigated previously, and was shown to cause alternative splicing at the 5’ end of exon 2, leading to the absence or presence of 9 amino acids in the signal peptide region ^43^. Due to the difficulty of determining full exon connectivity from short-read sequencing data, the distribution of isoforms remained unclear. Therefore, we have decided to use nanopore sequencing technology allowing us to investigate sequencing long reads. We obtained 12 stomach RNA samples from GTEx, six from homozygous carriers of the rs4072037 major allele T, and six from homozygous carriers of the minor allele C ^34^. We amplified the MUC1 region and sequenced all barcoded amplicons on a single flow cell on the MinION sequencing device. In total, we obtained 1,121 reads spanning the full-length MUC1 isoforms, i.e. having assignable barcodes at both ends for downstream analysis (**Figure S3 and S4; Table S2**). We exclusively observed an isoform with a short exon 2 in the TT individuals, whereas a longer isoform was always present in the CC individuals, thereby confirming the modulating effect of rs4072037 genotype on MUC1 splicing.

### Similar results using a case-control study design

We then investigated whether the genetic architecture of antibody levels against human polyomaviruses is similar to that of serostatus to these same polyomaviruses. Serological status was defined based on the manufactures defined cut-off of 250 MFI (see Methods). Individuals with levels of IgG antibodies above the threshold were considered seropositive (cases), while those below the threshold were seronegative (controls) (**Table 1**). Case-control GWAS using logistic regression were performed independently in the three cohorts and meta-analyzed ^32^. Q-Q plots revealed no inflation of the test statistics for HPyV6 (λ=0.99) and JCPyV (λ=0.98), while MCPyV (λ=0.90) and BKPyV (λ=0.72) showed varying degrees of deflation (**Figure S1**). Chicago plots for the genome-wide significant loci are presented in **Figure 2**. We observed again evidence of an involvement of HLA class II variation for JCPyV (rs9271025, P=4.31e-46) and MCPyV (rs73401330, P=8.49e-29) (**Table 2**). Strongest associations with seropositivity were observed for HLA-DQB1 allele. We also replicated the associations observed in *FUT2* for JCPyV (rs2432132, P=1.70e-15) and *STING1* for MCPyV (rs55792153, P=2.97e-09). Associations with *FUT2* and *HLA* variants were not replicated for BKPyV and HPyV6, respectively. In addition, the case-control analyses did not reveal any additional associated region, suggesting that there are no SNPs purely associated with susceptibility.

## Discussion

Human polyomaviruses are highly prevalent and can cause severe disease in immunocompromised individuals. They present a wide range of tissue tropisms and appear to cause neurologic, renal, and skin diseases ^48–50^. While their molecular mechanisms of infection are well known, the potential impact of human genetic variation on their lifecycle and pathogenicity remains understudied. For this study, we were interested in gaining insight into the genetic underpinning of the humoral immune responses to five polyomaviruses: BKPyV, JCPyV, HPyV6, MCPyV and WUPyV. Their seroprevalence was high in all three cohorts included in our study (55-96%), consistent with previously published surveys and confirming that human polyomavirus infections are common in the general population ^3,51^.

Genes within the HLA region are prime candidates for susceptibility to viral infection and regulation of the immune response. HLA has been associated with multiple infectious diseases, such as HIV, hepatitis B, hepatitis C, and tuberculosis ^52^. In this study, we confirm the involvement of HLA variation in the modulation of humoral immune response to two polyomaviruses, JCPyV and MCPyV ^3,5,53^. Specifically, we observed the strongest associations with quantitative antibody levels for HLA-DRB1 and HLA-DQA1 alleles, and with seropositivity for HLA-DQB1 allele.

Our study also shows that *FUT2* variation (rs601338 G>A) plays a role in the determination of antibody levels against BKPyV and JCPyV VP1. The FUT2 enzyme catalyzes the addition of a fucose residue to type 1 glycoprotein chains to form ABH blood group antigens that are released in secretions and at mucosal surfaces ^54^. Consistent with previous reports, 78% of our study population carried at least one functional allele, which is sufficient to render an individual FUT2 secretor, whereas 22% of individuals were found to be homozygous for the nonsense variant leading to defective FUT2 and the absence of ABH antigens in secretions (FUT2 non-secretors) ^33,55^. Interestingly, non-secretors had significantly higher IgG titers against BKPyV and JCPyV compared to secretors, suggesting higher antigen exposure and/or stronger immune response. One hypothetic explanation is that the binding of polyomaviruses to host cell sialic acid receptors could be impaired in the presence of FUT2-dependent blood antigens on the mucosal surfaces. Consequently, non-secretors might be more permissive to viral propagation and replication than secretors. The same *FUT2* variant is known to have an impact on several infections: non-secretors are protected against norovirus and rotavirus, but are at higher risk of infection with other pathogens, including e.g. *Candida albicans, Streptococcus pneumoniae* and mumps ^55–60^. The capacity to generate soluble ABH blood group antigens, dependent on the functionality of the FUT2 enzyme, represents an important modulator of infectious disease susceptibility in humans.

Our analyses confirm the implication of the *STING1* gene in the determination of antibody levels against MCPyV VP1, which was recently reported by Kachuri *et al*. ^61^. The variants most associated with anti-MCPyV IgG levels are strong regulators of the expression levels of *STING1*. The encoded protein, STING, plays a pivotal role in anti-viral innate immunity by activating type I interferons (IFN-I) and proinflammatory cytokines. Individuals carrying the homozygous genotype for the rs7444313 minor allele (GG) had significantly higher anti-MCPyV IgG levels. A recent study by Kennedy *et al*. reported an association between the same variant and the intensity of the immune response following primary smallpox vaccination ^62^. The same study also showed that expression of the correlated non-synonymous rs1131769 variant (R232H) in the *STING1* gene leads to a decrease in IFN-α expression levels. This result substantiates a previous finding showing reduced IFN-β transcription for carriers of the R232H variant ^63^. Evidence suggests that this amino acid change may have a modifying effect on the function of STING. Due to a weaker binding between cGAMP and the H232 variant, STING might be less effective, resulting in a lower immune response. In turn, this might allow higher viral replication resulting in increased anti-MCPyV IgG levels. Interestingly, rs7444313 has significantly different allele frequencies in European populations (ref. allele: G=0.28) relative to African populations (ref. allele: G=0.83), suggesting a possible role of infection-driven selection in this genomic region.

We observed a very strong association signal with anti-WUPyV IgG response in *MUC1*, the gene encoding the Mucin1 protein. There is no established role of WUPyV in disease thus far. However, it was detected in the trachea of immunocompromised children, in close proximity to MUC5AC expressing cells ^64^. Physiologically, cell surface mucins form a network that contributes to the mucosal barrier to infection ^65^. MUC1, in particular, consists of a large extracellular O-glycosylated polypeptide backbone that extends 200-500 nm above the apical surface ^66^. The minor allele (T) of the top associated SNP, rs4072037, which associates with higher IgG levels, is known to alter the physiological functions of MUC1 because it leads to an alternative splicing of the 5’-region of exon 2. As a consequence, a shorter protein isoform is produced, which impairs the protective function of MUC1 on the gastric mucosa ^67^. The rs4072037 T/T genotype has been previously associated with an increased risk of gastric cancer ^67,68^, most likely due to a concomitant increase in the risk of gastritis due to *Helicobacter pylori* infection ^69^. The newly discovered association with humoral response to WUPyV is most likely coincidental. Indeed, the absence of any trace of WUPyV in cancer samples suggests that this virus is unlikely to play any role in gastric carcinogenesis ^70^. However, our results suggest once again that MUC1 plays an essential role in the protection of gastric mucosa, and that a common, genetically-encoded alteration of MUC1 has a deleterious impact on that important physiological function.

For human polyomaviruses with seroprevalence below 95%, we investigated the relationship between the genetic contribution to serostatus and humoral immune response in seropositive individuals. Whereas the quantitative approach is purely looking at immune response in infected individuals, the case-control study design has the potential to also uncover genetic loci involved in susceptibility to infection. Our meta-analysis results suggest very similar genetic contributions to the continuous (quantitative) and binary (case-control) phenotypes, with comparable statistical strength. The same variable genetic loci in HLA class II, *FUT2* and *STING1* were found to be associated with JCPyV6 and MCPyV serostatus and IgG levels. These similarities in the results could be explained by the impossibility of being able to differentiate between individuals who were never infected and individuals who were infected but had an extremely weak antibody response.

Here we report the first meta-analyses aiming to identify human genetic determinants of the humoral immune response against multiple polyomaviruses. Statistically combining data from CoLaus, GRAS and a subset of the UKB, allowed us to run analyses on a total of 15,660 individuals of European ancestry. This provided greater statistical power and a more accurate estimate of the underlying effects, also reducing the risk of false-negative results. Some limitations need however to be considered. First, we did not have serological data for all five polyomaviruses in each cohort. Second, the meta-analyses relied on summary statistics derived from independently computed GWAS, which might result in biases due to differences in the design, analysis and conduct of the individual studies. Third, our analyses were restricted to individuals of European ancestry, which limits the generalizability of our results. Finally, as mentioned above, individuals with very low IgG levels were excluded from the quantitative association analyses, due to the impossibility of distinguishing them from truly seronegative (i.e. never infected) study participants; this resulted in reduced statistical power.

In summary, we here report strong associations of *HLA* class II, *FUT2, STING1*, and *MUC1* genetic variants with the intensity of the humoral IgG response to multiple human polyomaviruses. Together, these results demonstrate the modulating contribution of host genetic variation to the individual response against some of the most prevalent human viruses.

## Data Availability

Full meta-analyses association results are available for download from Zenodo.

https://doi.org/10.5281/zenodo.4189621

