## Supplementary Tables and Figures for "Human genomics of the humoral immune response against polyomaviruses"

**Table S1.** Results from the serological analyses of rs4072037 for six different *H. pylori* and four EBV antigens in CoLaus study.

| Pathogen | Antigen | Prevalence (%) | Beta | SE | P-value |
|---|---|---|---|---|---|
| **EBV** | VCA p18 | 93 | 7.1e-03 | 2.3e-02 | 7.6e-01 |
| | EBNA | 90 | 9.6e-03 | 2.3e-02 | 6.8e-01 |
| | Zebra | 88 | -1.9e-03 | 2.3e-02 | 9.3e-01 |
| | EA-D | 77 | 4.2e-02 | 2.3e-02 | 7.0e-02 |
| ***H. pylori*** | HP1564 OMP | 28 | -2.0e-02 | 2.3e-02 | 3.8e-01 |
| | HP10 GroEL | 27 | -8.5e-03 | 2.2e-02 | 7.1e-01 |
| | HP547 CagA | 17 | -9.7e-03 | 2.3e-02 | 6.7e-01 |
| | HP887 VacA | 17 | 2.5e-02 | 2.3e-02 | 2.7e-01 |
| | HP73 UreaseA | 15 | -1.8e-02 | 2.3e-02 | 4.3e-01 |
| | HP875 Catalase | 14 | -1.4e-03 | 2.3e-02 | 9.5e-01 |
| **WUPyV** | VP1 | 96 | -2.5e-01 | 2.3e-02 | 6.2e-28 |

**Table S2. Read counts, as mapped to the different *MUC1* isoforms.**

| Genotype | Sample | NM_001018017 | NM_002456 | NM_001018016 | NM_001204287 | Total |
|---|---|---|---|---|---|---|
| **CC** | **1** | 67 | 7 | 0 | 0 | 74 |
| **CC** | **2** | 69 | 14 | 0 | 0 | 83 |
| **CC** | **3** | 30 | 3 | 0 | 0 | 33 |
| **CC** | **4** | 85 | 19 | 0 | 0 | 104 |
| **CC** | **5** | 105 | 27 | 0 | 0 | 132 |
| **CC** | **6** | 91 | 19 | 0 | 0 | 110 |
| **TT** | **7** | 0 | 0 | 74 | 37 | 111 |
| **TT** | **8** | 0 | 0 | 80 | 23 | 103 |
| **TT** | **9** | 0 | 0 | 63 | 17 | 80 |
| **TT** | **10** | 0 | 0 | 87 | 20 | 107 |
| **TT** | **12** | 0 | 0 | 77 | 20 | 97 |
| **TT** | **12** | 0 | 0 | 70 | 17 | 87 |
| **Total** | | 447 | 89 | 451 | 134 | **1121** |

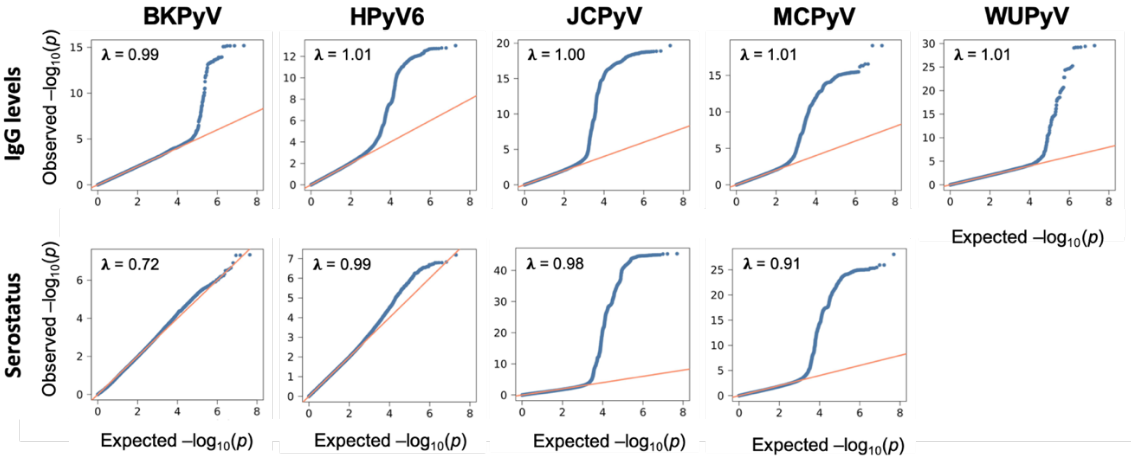


**Figure S1. Quantile-Quantile (Q-Q) plots of SNPs for association to IgG levels and serostatus in the meta-analysis of CoLaus, UKB and GRAS.** On the y-axis, the observed p-values (blue dots) are plotted against the expected p-values under the null distribution. The red line indicates the distribution of SNPs under the null distribution. Lambda (λ) denotes the genomic control inflation factor.

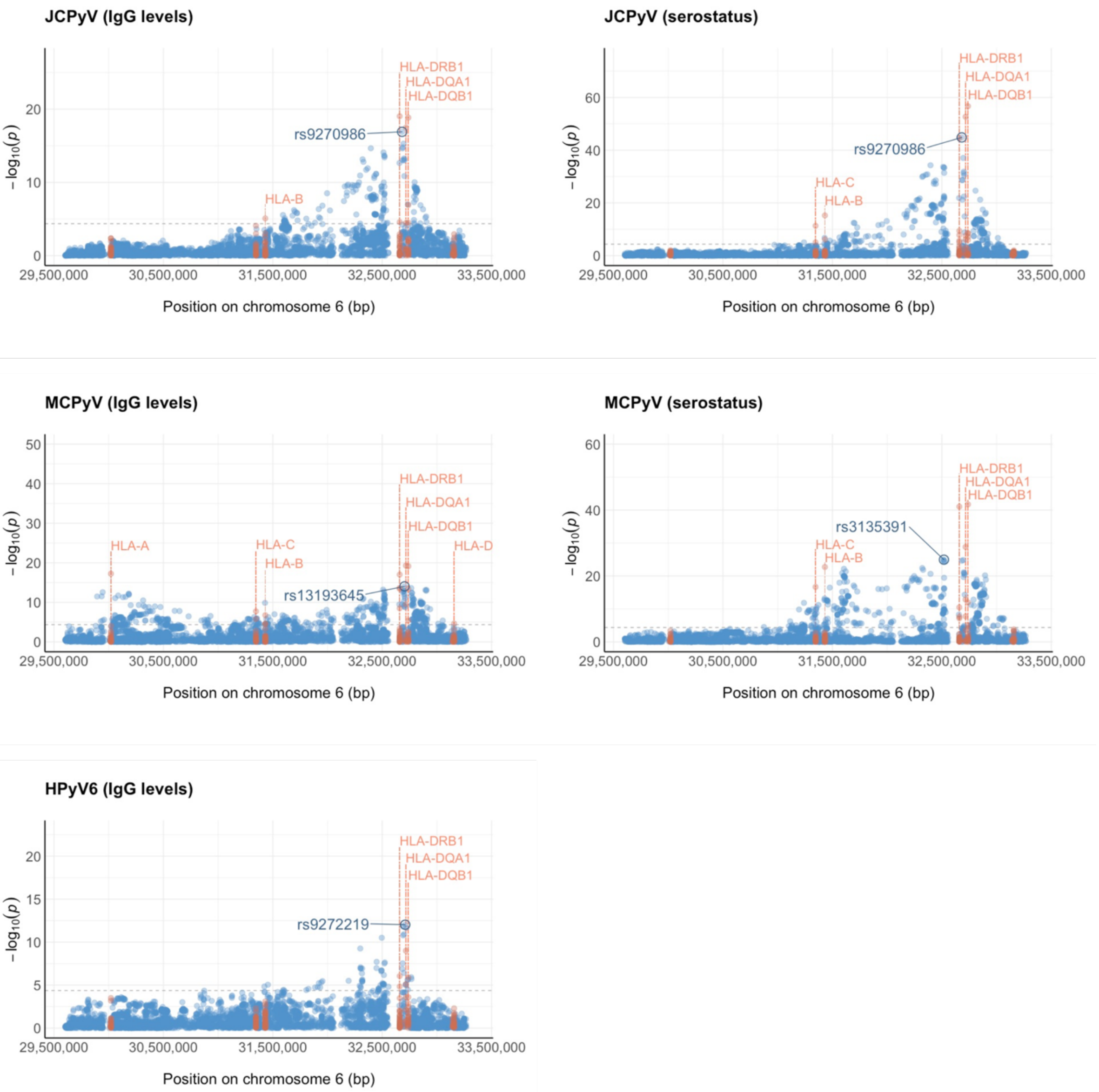


**Figure S2. Regional association plot of SNP2HLA results for BKPyV, JCPyV and MCPyV.** For each SNP (blue) and each classical HLA allele (orange), the P-value (in $-\log_{10}$) is plotted against its position in the MHC genomic region on chromosome 6 (UCSC hg17, NCBI Build 35). The most significant association was observed for rs9270986. The dashed horizontal line indicates the threshold for HLA-wide significance (P=4.3e-05). The annotated dashed orange vertical lines indicate the positions of the significant HLA alleles. The circled point represents the top associated SNP.

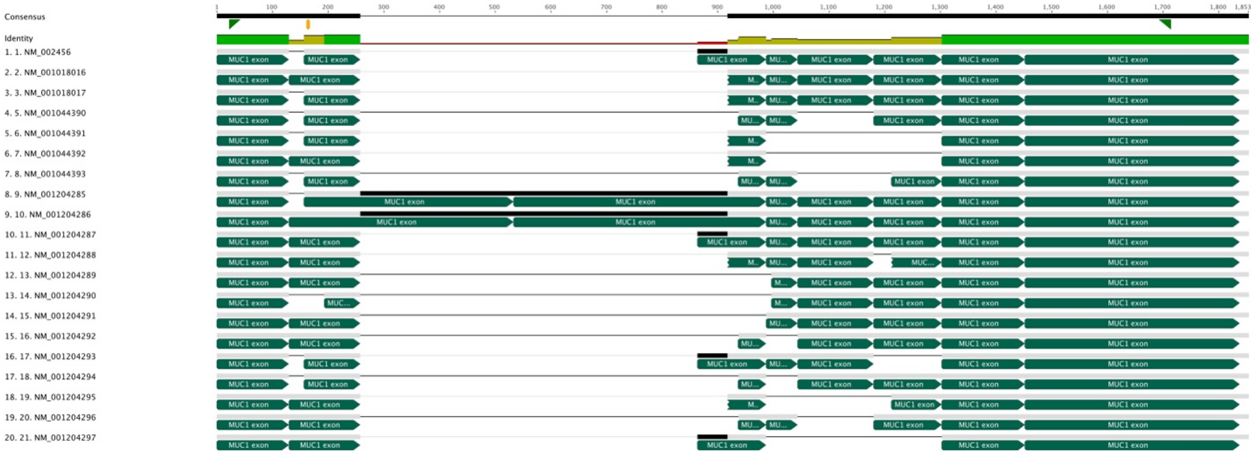


**Figure S3. Structural representation of *MUC1* isoforms.** The exons are represented with green boxes.

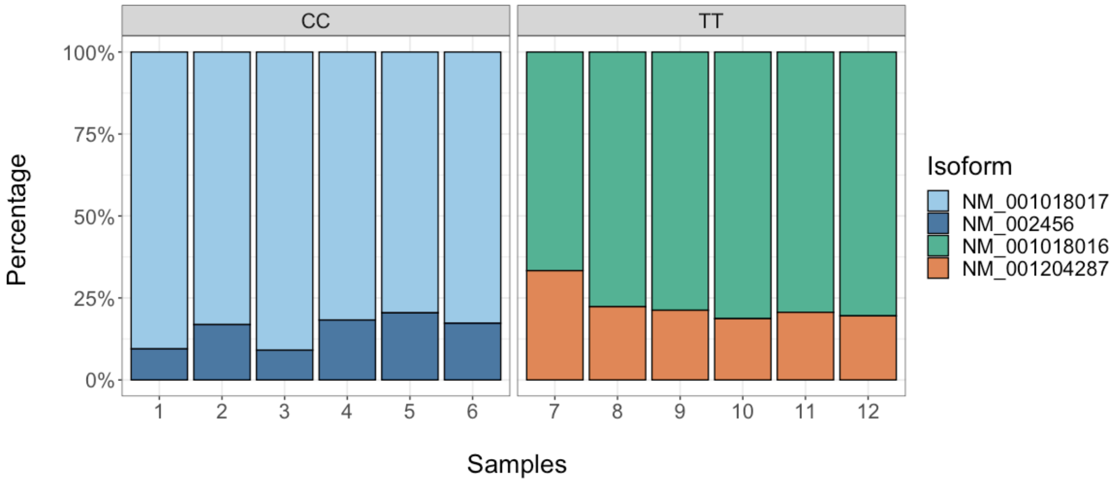


**Figure S4. Percentage of reads, as mapped to the different *MUC1* isoforms.** The plot shows the percentage of mapped reads for the 12 stomach RNA samples from GTEx, 6 of which from homozygous carriers of the rs4072037 major allele T (left panel), and 6 from homozygous carriers of the minor allele C (right panel).
